# The intermediary effect of emotional exhaustion on the relationship between moral dilemmas and turnover intention among psychiatric nurses

**DOI:** 10.1101/2025.11.06.25339703

**Authors:** Jiehui Yang, Rong Zhang, Yunling Zhong, Yana Wang, Shengyu Liu

**Affiliations:** School of Nursing, Guangdong Pharmaceutical University, Guangzhou, Guangdong, China; Office of the President, Guangdong Second Rongjun Youfu Hospital, Foshan,Guangdong, China

## Abstract

Psychiatric nurses frequently encounter moral dilemmas and emotional exhaustion, both of which significantly influence job satisfaction and turnover intentions. Studies highlight that these factors are key drivers of nurse attrition. Given the high turnover rates in this field, understanding the mechanisms connecting these factors is essential.The aim of this study was to investigate the current status of moral dilemmas, emotional exhaustion, and turnover intentions among psychiatric nurses, and to explore the mediating effect of emotional exhaustion in the relationship between moral dilemmas and turnover intentions.A convenience sampling method was used to select psychiatric nurses from four secondary and tertiary hospitals in Guangdong Province as the study subjects in February 2025. Data were collected using a descriptive information form, the Moral Dilemmas Scale for Psychiatric Nurses, the Emotional Exhaustion Scale, and the Turnover Intention Scale. Path analysis was conducted using structural equation modeling with AMOS 24.0, and the mediating effect was tested.A total of 403 nurses participated in the study. The mean score for moral dilemmas was (30.10 ± 32.20), for emotional exhaustion was (12.45 ± 6.68), and for turnover intention was (2.37 ± 0.94). The total score of moral dilemmas was positively correlated with the total score of emotional exhaustion (*r* = 0.400, *p* < 0.01), the total score of emotional exhaustion was positively correlated with the total score of turnover intention (*r* = 0.389, *p*< 0.01), and the total score of moral dilemmas was positively correlated with the total score of turnover intention (*r* = 0.514,*p* < 0.01). Emotional exhaustion partially mediated the relationship between moral dilemmas and turnover intention, with a mediating effect value of 0.172, accounting for 42.86% of the total effect. Emotional exhaustion can be considered as a key mediator in the relationship between moral dilemmas and turnover intentions in psychiatric nursing.Moral dilemmas and turnover intentions were at a moderate to low level, and emotional exhaustion was at a moderate level for this group of nurses. Emotional exhaustion has an effect on the relationship between moral dilemmas and turnover intentions among psychiatric nurses.

## Introduction

The shortage of nursing human resources has become a major challenge for healthcare systems worldwide, and nurse turnover is one of the primary causes of this shortage [1]. Nurses’ turnover intention refers to their propensity to resign from their current job or unit, and this intention is an effective predictor of actual turnover behavior [2]. Studies have found that between 20.2% and 56.1% of nurses in China have a high turnover intention [3], with psychiatric nurses exhibiting a particularly high rate of 70.2%, due to factors such as workplace violence and moral dilemmas [4,5]. This phenomenon has severely threatened the stability of psychiatric nursing services.

Moral dilemma refers to the psychological distress experienced by nurses who, when confronted with moral choices, are unable to act in accordance with their perceived moral norms due to personal or external constraints [7]. Prolonged exposure to moral dilemmas has been shown to lead to job fatigue and increased occupational stress, which in turn triggers burnout and exacerbates turnover intention [8].

Emotional exhaustion refers to the depletion of an individual’s emotional and psychological resources under sustained work pressure, leading to fatigue, energy depletion, and a loss of enthusiasm for work [9]. Studies have demonstrated a close correlation between emotional exhaustion and turnover intention. As work stress increases, emotional exhaustion rises, which further contributes to turnover intention [10].

According to Resource Conservation Theory, individuals exposed to persistent stressors without sufficient resources to cope with them experience a loss of psychological and emotional resources, leading to emotional exhaustion [11]. This exhaustion, in turn, becomes a key factor that increases turnover intention. Prolonged emotional exhaustion can cause burnout and prompt individuals to avoid their jobs, ultimately leading to thoughts of leaving and seeking alternative employment.

Psychiatric nurses, in particular, expend significant emotional resources when faced with moral dilemmas. If these resources are not replenished, they are prone to emotional exhaustion [12]. In a state of emotional depletion, individuals may consider leaving their jobs because they feel unable to cope with the ongoing work pressures [13,14].

Thus, there is an association between moral dilemmas, emotional exhaustion, and turnover intention among psychiatric nurses. However, the relationship among these factors, as well as the mechanism through which moral dilemmas influence turnover intention, remains unclear. Most domestic and international studies have focused on the relationship between job stress and turnover intention, with fewer investigating the mediating role of emotional exhaustion, particularly within the group of psychiatric nurses. Therefore, this study aims to explore the mediating role of emotional exhaustion between moral dilemmas and turnover intention, with the goal of providing effective intervention strategies for hospital administrators to reduce psychiatric nurses’ turnover intention.

## Methods

### Study design

This study adopted a cross-sectional design to investigate the mediating role of emotional exhaustion in the relationship between moral dilemmas and turnover intention among psychiatric nurses. The research was conducted in February 2025, using a convenience sampling method to select psychiatric nurses.

### Sample and participants

The study targeted 403 psychiatric nurses from four secondary or higher-level hospitals in Guangdong Province. The inclusion criteria were as follows: (1) possession of a valid nursing license, (2) registration as a nurse engaged in psychiatric nursing, and (3) voluntary participation with informed consent. The exclusion criteria included: (1) a history of psychiatric disorders or consciousness disturbances, and (2) not being on duty during the study period (e.g., on leave, attending training, or studies).

Structural equation modeling (SEM) will be used for data analysis, with a required sample size of no less than 200 participants[15]. According to the sample size estimation method proposed by Kendall, the sample size should be 10 to 20 times the number of variables[16]. Given that the study involves 15 variables, the estimated sample size is between 150 and 300. Taking into account a 20% attrition rate, the minimum required sample size is 360 participants.

A total of 425 questionnaires were distributed. The responses were double-checked and invalid questionnaires were excluded based on the following criteria, drawn from previous studies: (1) response time < 2 minutes, (2) incorrect or missing answers, and (3) more than 80% of consecutive answers being identical. As a result, 403 valid questionnaires were retrieved, yielding a response rate of 94.8%.

### Data collection

In this study, the questionnaire was distributed via the Questionnaire Star platform. Participants were required to read the informed consent form before completing the survey and could only proceed after agreeing to the terms. The purpose and significance of the study were outlined on the first page of the questionnaire. Participants filled out the survey anonymously, and all questions had to be answered before submission. To prevent duplicate responses, each IP address was allowed to submit the questionnaire only once.

### Measurements

#### Descriptive information form

This form was designed by the researchers based on relevant literature and consisted of 10 questions, which included gender, age, years of experience in psychiatry, average monthly income, hospital grade, marital status, education level, professional title, position, and nature of work[6, 17].

#### Moral Dilemma Scale for Psychiatric Nurses

The scale was developed by Liu Xiaopei et al. and consists of three dimensions: low autonomy, failure to uphold the patient’s best interests, and value conflict, with a total of 20 items[18] . Each item is divided into two parts: frequency of occurrence (0 = never, 1 = occasionally, 2 = sometimes, 3 = frequently, 4 = very frequently) and degree of distress (0 = none, 1 = mild, 2 = moderate, 3 = severe, 4 = very severe), rated using a 5-point Likert scale. The total score is the sum of the products of the frequency and degree of distress for each item, with a score range of 0 to 320. Higher total scores indicate more severe moral dilemmas faced by psychiatric nurses. The Cronbach’s alpha coefficient for the scale was 0.940, with the Cronbach’s alpha coefficients for the dimensions ranging from 0.705 to 0.927. In this study, the Cronbach’s alpha coefficient for the scale was 0.955.

#### Emotional Exhaustion Scale

The scale was derived from the Emotional Exhaustion subscale of the Burnout Scale revised by Li Chaoping and Shi Kan [19], which is used to assess the degree of depletion of an individual’s emotional resources. The scale consists of five items and is rated on a 7-point Likert scale: (0 = “very incongruent,” 1 = “incongruent,” 2 = “somewhat incongruent,” 3 = “neutral,” 4 = “somewhat congruent,” 5 = “congruent,” 6 = “very congruent,” and 7 = “extremely congruent”). The total score ranges from 0 to 30, with higher scores indicating more severe emotional exhaustion. The Cronbach’s alpha coefficient for this scale is 0.88, and in this study, it was 0.937.

#### Turnover Intention Scale

The scale was developed by Spector et al. in 1988 and consists of a single item designed to assess employees’ intention to leave[20]. The scale uses a 5-point Likert scale, with scores ranging from 1 to 5, representing “never” to “extremely frequently,” respectively. Higher scores indicate a stronger intention to leave. The Cronbach’s alpha coefficient for the scale is 0.83.

#### Data analysis

Data analysis in this study was conducted using SPSS Statistics 27.0 (Statistical Package for the Social Sciences) statistical software. Categorical data were presented as frequencies and proportions, while normally distributed continuous data were expressed as mean ± standard deviation. Pearson correlation analysis was used to examine the relationships between psychiatric nurses’ moral dilemmas, emotional exhaustion, and turnover intention. Structural equation modeling was performed using AMOS 24.0 (Analysis of Moment Structures), and the significance of the mediating effect was tested using the Bootstrap method, with a significance level set at α= 0.05.

#### Ethics statement

Ethics committee approval for the research was obtained from the Ethics Committee of Guangdong Second Rongjun Youfu Hospital (Approval No:

AF/SC-01/01.0-20), and institutional permission was obtained from the Guangdong Second Rongjun Youfu Hospital. Participants were informed about the aim of the research, and they were told that their participation was voluntary. Their consent was obtained through an informed consent form.

## Results

### Sample characteristics

The study contacted 403 participants, and their socio-demographic characteristics are presented in Table 1.

**Table 1.** Socio-demographic characteristics of psychiatric nurses(*n*=403).

| Socio-demographic characteristics | n | % |
| --- | --- | --- |
| Gender |  |  |
| Male | 75 | 18.6 |
| Female | 328 | 81.4 |
| Age (years) |  |  |
| 20-25 | 192 | 47.6 |
| 26-30 | 105 | 26.1 |
| 31-40 | 80 | 19.9 |
| >40 | 26 | 6.5 |
| Marital status |  |  |
| Unmarried | 258 | 64.0 |
| Married | 141 | 35.0 |
| Divorced or Widowed | 4 | 1.0 |
| Professional title |  |  |
| Registered Nurse | 203 | 50.4 |
| Nurse practitioner | 125 | 31.0 |
| Charge Nurse | 66 | 16.4 |
| Assistant Head Nurse or Nurse Manager | 9 | 2.2 |
| Years of Work Experience(years) |  |  |
| 1-5 | 283 | 70.2 |
| 6-10 | 62 | 15.4 |
| 10-20 | 45 | 11.2 |
| >21 | 13 | 3.2 |
| Education level |  |  |
| Associate Degree or Below | 134 | 33.3 |
| Bachelor's Degree | 248 | 61.5 |
| Master's degree or above | 21 | 5.2 |
| Position |  |  |
| Clinical nurse | 349 | 86.6 |
| Nursing Team Leader | 26 | 6.5 |
| Nursing manager | 28 | 6.9 |
| Hospital Grade |  |  |
| Secondary hospital | 201 | 49.9 |
| Tertiary Hospital | 202 | 50.1 |
| Employment Method |  |  |
| Compilation | 138 | 34.2 |
| Contract employment | 221 | 54.8 |
| Labour dispatch | 44 | 10.9 |
| Average monthly income (RMB) |  |  |
| <5000 | 145 | 36.0 |
| 5000-8000 | 204 | 50.6 |
| 8000-12000 | 41 | 10.2 |
| >12000 | 13 | 3.2 |

Values are n (%). Percentages are calculated on the total sample . Professional title and work experience are grouped as shown; hospital grade refers to secondary and tertiary hospitals in Guangdong Province.

### The Current Situation of Moral dilemma, Emotional Exhaustion, and Turnover Intention in Psychiatric Nurses

The overall score of moral dilemma among psychiatric nurses was (30.10±32.20), with an average item score of (1.51±1.61). Across dimensions, the item scores ranked in descending order as follows: value conflict (1.99±2.15), insufficient autonomy (1.43±1.66), and not maintaining the patient’s best interests (1.41±1.58).Emotional exhaustion had a total score of (12.45±6.68), with a mean item score of (2.49±1.34).The score for turnover intention was (2.37±0.94).

### Correlational Analysis of Moral dilemma, Emotional Exhaustion, and Turnover Intention in Psychiatric Nurses

Pearson correlation analysis indicated that Moral dilemma showed a positive correlation with emotional exhaustion (*r*=0.400, *P*<0.01), Moral dilemma was positively associated with turnover intention (*r*=0.389, *P*<0.01), and emotional exhaustion was positively associated with turnover intention (*r*=0.514, *P*<0.01(Table 2).

**Table 2.** Correlation Analysis of Moral dilemma, Emotional Exhaustion, and Turnover Intention in Psychiatric Nurses (*n*=403).

| Variables | Moral dilemma | Insufficient autonomy | Failure to uphold patients' best interests | Value conflict | Emotional exhaustion | Turnover intention |
| --- | --- | --- | --- | --- | --- | --- |
| Moral dilemma | 1 |  |  |  |  |  |
| Insufficient autonomy | 0.978** | 1 |  |  |  |  |
| Failure to uphold patients' best interests | 0.934** | 0.871** | 1 |  |  |  |
| Value conflict | 0.854** | 0.776** | 0.729** | 1 |  |  |
| Emotional exhaustion | 0.400** | 0.376** | 0.398** | 0.349** | 1 |  |
| Turnover intention | 0.389** | 0.385** | 0.368** | 0.310** | 0.514** | 1 |

Entries are Pearson correlation coefficients (two-tailed). Moral dilemma subscales are Insufficient autonomy, Failure to uphold patients' best interests, and Value conflict. Diagonal values are 1. Higher scores indicate higher levels of each construct. \*\* $p<.01$ .

### Mediation analysis of emotional exhaustion

#### Regression analysis of the influence of Moral dilemma and emotional exhaustion on turnover intention in psychiatric nurses

Using the Moral dilemma score and emotional exhaustion score, which were statistically significant in the correlation analysis, as independent variables and the turnover intention score as the dependent variable, multiple linear regression analysis was conducted.The results showed that both Moral dilemma and emotional exhaustion entered the regression equation, jointly explaining 30.0% of the total variance in turnover intention among psychiatric nurses.Moral dilemma positively influenced turnover intention, and emotional exhaustion positively predicted turnover intention (Table 3).

**Table 3.**
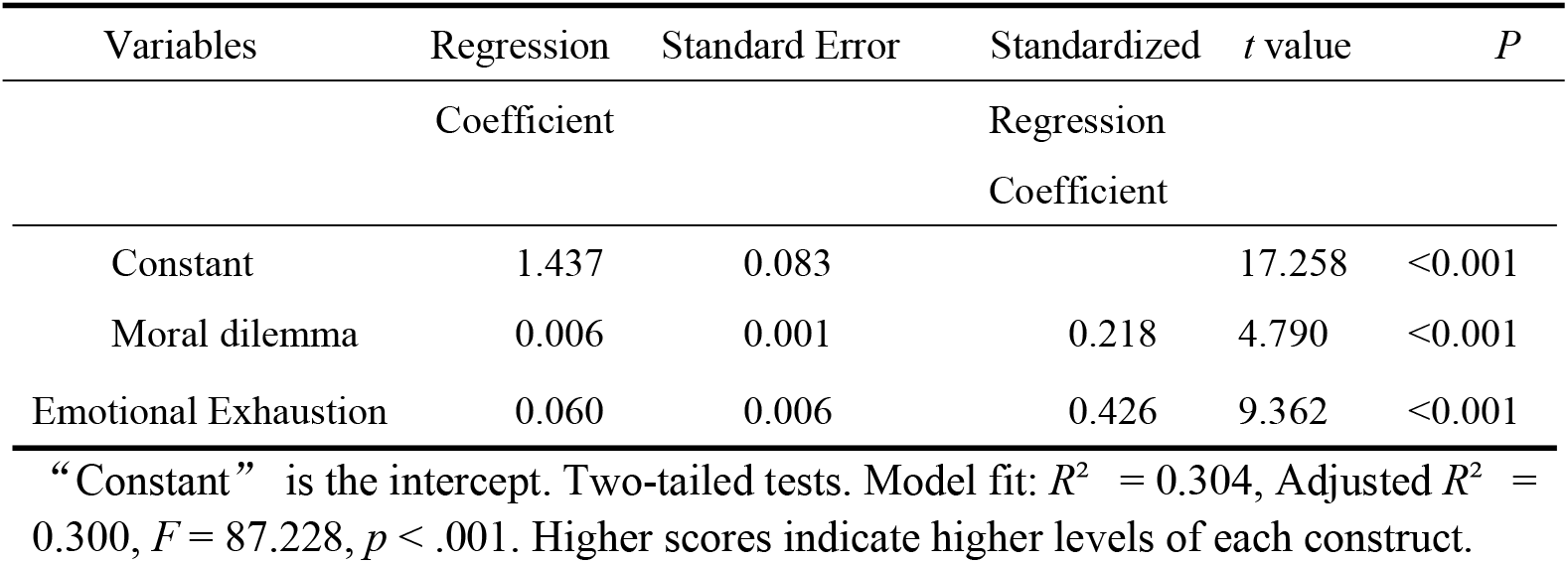
Regression analysis of the influence of Moral dilemma and emotional exhaustion on turnover intention among psychiatric nurses (*n*=403).

#### Mediation analysis of emotional exhaustion between Moral dilemma and turnover intention in psychiatric nurses

Structural equation modeling was constructed using AMOS 24.0 software, with Moral dilemma as the independent variable, emotional exhaustion as the mediating variable, and turnover intention as the dependent variable. Maximum likelihood estimation was used to fit the model.The fit indices include the chi-square/df ratio (χ 2 / df), goodness-of-fit index (GFI), adjusted goodness-of-fit index (AGFI), normed fit index (NFI), non-normed fit index (TLI), comparative fit index (CFI), incremental fit index (IFI), and root mean square error of approximation (RMSEA).The results showed that the model fit indices were satisfactory(Table 4).Finally, a mediating effect model of emotional exhaustion between Moral dilemma and turnover intention among psychiatric nurses was constructed(Fig 1).

**Table 4.**
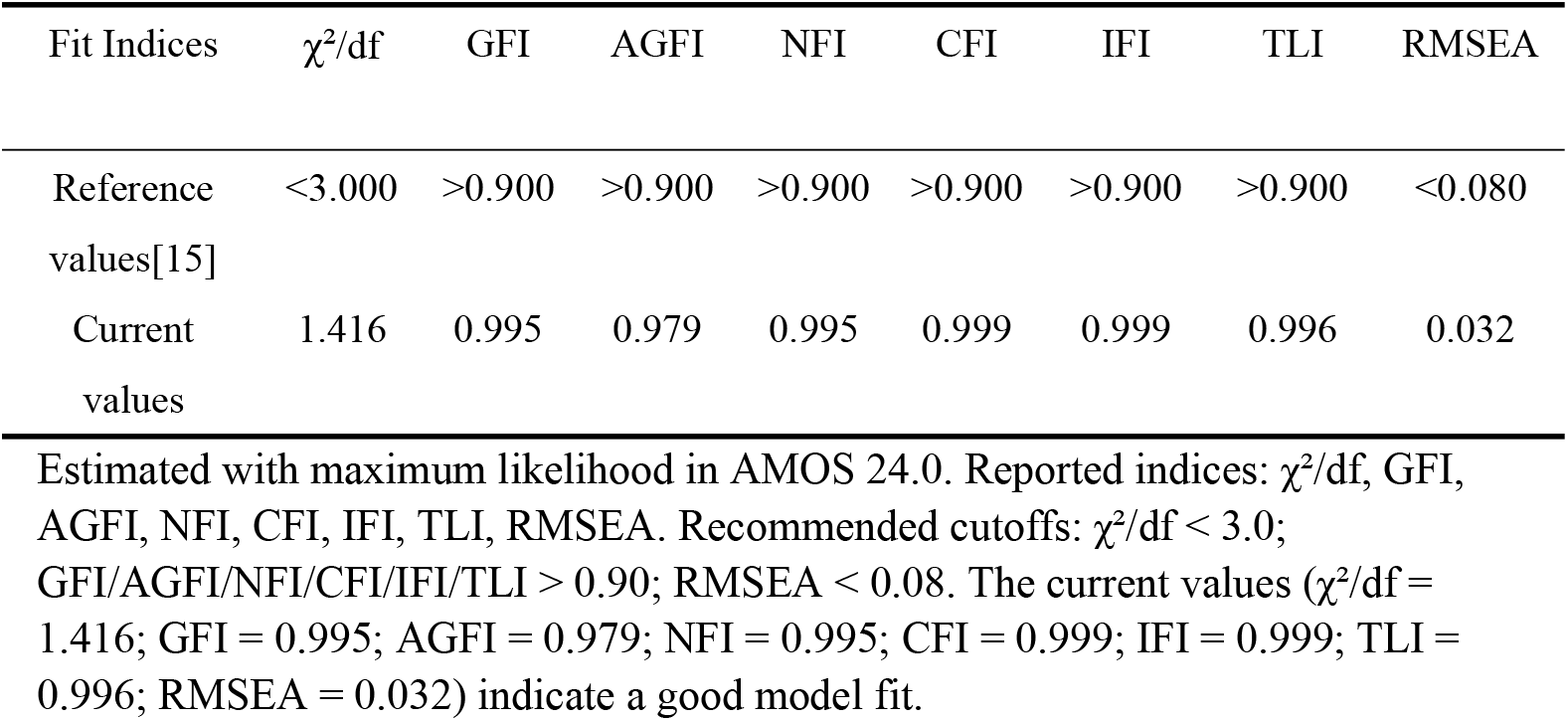
Model Fit Indices for Overall Model Evaluation.

**Fig 1.** Structural equation modeling of the relationships among Moral dilemma, emotional exhaustion, and turnover intention in psychiatric nurses. Moral distress (latent construct derived from moral dilemma dimensions) is indicated by Insufficient autonomy, Failure to uphold patients’ best interests, and Value conflict (residuals e1–e3). Residuals for Emotional exhaustion and Turnover intention are e4 and e5. Numbers on arrows are standardized coefficients: Moral distress → Emotional exhaustion *β* = 0.41; Emotional exhaustion → Turnover intention *β* = 0.42; Moral distress → Turnover intention β = 0.23; loadings to Moral distress = 0.96, 0.91, 0.81. All paths *p* < .01.Indirect effect of moral distress on turnover intention via emotional exhaustion = 0.172 (42.86% of total effect).

To examine the mediation effects in the model, the Bootstrap method was employed with 5000 bootstrap resamples and a 95% confidence interval.Results demonstrated that Moral dilemma was a significant predictor of emotional exhaustion (a = 0.409, SE = 0.018, *P* < 0.001).With both emotional exhaustion and Moral dilemma entered into the regression equation, emotional exhaustion significantly predicted turnover intention (b = 0.420, SE = 0.006, *P* < 0.001).Moral dilemma also significantly predicted turnover intention (c’ = 0.229, SE = 0.003, *P* < 0.001).The 95% confidence intervals for both the direct and indirect effects from Moral dilemma to turnover intention excluded zero, indicating that the mediation model via emotional exhaustion was supported.The indirect effect accounted for 42.86% (ab/(ab+c’)) of the total effect, with detailed results presented (Table 5).

**Table 5.** Mediation Effect Analysis of Emotional Exhaustion.

| Moral dilemma → Turnover intention | Estimated coefficient<br>t | Coefficient product |  | 95% bias-corrected bootstrap confidence interval |  | P |
| --- | --- | --- | --- | --- | --- | --- |
|  |  | Standard error | Z value | Lower limit | Upper limit |  |
| Total effect | 0.401 | 0.048 | 8.354 | 0.305 | 0.490 | <0.001 |
| Direct effect | 0.229 | 0.053 | 4.320 | 0.007 | 0.019 | <0.001 |
| Indirect effect | 0.172 | 0.031 | 5.548 | 0.113 | 0.233 | <0.001 |

Standardized total, direct, and indirect effects are reported. Indirect effect was tested with the product-of-coefficients and bias-corrected bootstrap 95% CIs (5,000 resamples); significance is inferred when the CI excludes 0 (all $p < .001$ ). $Z =$ coefficient/SE. Indirect effect = 0.172, accounting for 42.86% of the total effect.

## Discussion

### Current status of moral dilemma, emotional exhaustion, and turnover intention among psychiatric nurses

The present study found that the total score for moral dilemma among psychiatric nurses was (30.10±32.20), which, when compared with the midpoint of the scale (160.00), reflected a mild level of moral dilemma. This score was lower than that reported by Liu Xiaopei [18] (50.42±42.802). The discrepancy may be attributed to the timing of Liu’s data collection, which coincided with the COVID-19 pandemic. During that period, nurses not only had to adhere to routine hospital policies but also faced additional pressures from infection control requirements. It is also possible that, in recent years, the growing research attention on nurses’ moral dilemma has led to enhanced ethical support and training programs [21].The “value conflict” dimension yielded the highest score, consistent with the findings of Liu Xiaopei [18] and Xue B [22]. This suggests that psychiatric nurses frequently encounter value-based conflicts—having to make difficult decisions between respecting patients’ autonomy and safeguarding their welfare, or between upholding professional ethics and responding to patients’ needs [22].These conflicts add to nurses’ psychological strain, diminish job satisfaction, and may ultimately contribute to a stronger intention to leave.These findings highlight the need for nursing administrators to focus more on the psychological and emotional support of psychiatric nurses confronting value conflicts. Implementing comprehensive ethical training, clear moral guidance, and accessible psychological support systems can assist nurses in making appropriate decisions in complex situations, thereby reducing the stress arising from value conflicts.

The total score for emotional exhaustion among psychiatric nurses was (12.45±6.68), and the mean item score was (2.49±1.34). Compared to the theoretical midpoint of 3 for the scale items, this reflects a moderate level of emotional exhaustion. These findings align with the results of He Qing Sen [23], highlighting that emotional exhaustion among psychiatric nurses remains a significant concern.Psychiatric nurses face a unique work environment characterized by patients’ emotional instability, communication difficulties, and frequent emergencies, which increase the pressure on their management and response abilities. Furthermore, psychiatric patients often lack family support, and nurses are responsible for the overall safety and rehabilitation of these patients. Prolonged exposure to such high-pressure situations can lead to emotional exhaustion [24].According to the job demands-resources model, job demands refer to aspects of work that require continuous physical, mental, and emotional effort. Prolonged work demands can exacerbate emotional exhaustion. On the other hand, job resources, such as salary, supportive work environments, and opportunities for career development, help alleviate emotional exhaustion and promote personal growth [25].This suggests that administrators should carefully manage nurses’ work schedules, offer appropriate compensation, and foster a supportive work environment with ample opportunities for career growth.

The turnover intention score among psychiatric nurses was (2.37±0.94), with an average score of approximately 2.37. This indicates that the turnover intention of psychiatric nurses is at a moderately low level, close to “occasional” intent to leave. This is consistent with the findings of He Xuchong et al. [26].Psychiatric patients frequently exhibit symptoms like hallucinations and delusions, leading to verbal and physical aggression towards nurses. However, with the development and implementation of violence prevention training systems, the occurrence of violent incidents has decreased, helping to alleviate nurses’ negative emotions and effectively reduce their turnover intention [4].Currently, the employment market is highly competitive, making it more difficult to find a job. For re-employed nurses, their previous work experience and environment make them more sensitive to changes in roles and environments than newly graduated nurses. They may even struggle to adapt to new work environments. As a result, many nurses tend to retain their current positions when faced with employment pressure [27].This suggests that administrators should pay attention to the work environment of psychiatric nurses, continue to strengthen violence prevention training and psychological support, reduce negative emotions in the workplace, and thus lower turnover intention.

### The correlation between moral dilemma, emotional exhaustion, and turnover intention among psychiatric nurses

This study shows that moral dilemma among psychiatric nurses is positively correlated with emotional exhaustion, and moral dilemma is positively correlated with turnover intention. Emotional exhaustion is also positively correlated with turnover intention. Specifically, the higher the level of moral dilemma among psychiatric nurses, the higher the degree of emotional exhaustion; the higher the level of Moral dilemma, the higher the turnover intention; the higher the degree of emotional exhaustion, the higher the turnover intention. These results are consistent with those of Maunder RG [14].Psychiatric nurses frequently encounter moral dilemma in their work, particularly when making decisions about treatment plans, patient autonomy, and safeguarding patient safety. These challenges create substantial psychological pressure.Additionally, the multiple expectations placed on nurses by patients, colleagues, and management contribute to emotional exhaustion and psychological fatigue, thereby increasing turnover intention [28].As a result, healthcare organizations should establish ethics committees, develop a three-tiered prevention strategy, and formulate ethical guidelines to support the physical and mental health of nurses [28]. Nurses should regularly participate in discussions and analyses of ethical issues, encouraging them to voice their personal perspectives. The “4A”cycle model should be employed to assist nurses in alleviating moral dilemma. Departments should organize regular training sessions on Moral dilemma and reflective education to teach basic knowledge and coping strategies, thereby improving nurses’ ethical decision-making abilities [29].

### Emotional exhaustion plays a partial mediating role between moral dilemma and turnover intention among psychiatric nurses

The mediation analysis results suggest that emotional exhaustion partially mediates the relationship between moral dilemmas and turnover intention among psychiatric nurses.Specifically, both moral dilemmas (*β*=0.409, *P*<0.001) and emotional exhaustion (*β*=0.420, *P*<0.001) positively predict nurses’ turnover intention.Furthermore, moral dilemmas indirectly influence turnover intention through emotional exhaustion (*β*=0.229, *P*<0.001), with the mediating effect accounting for 42.86% of the total effect.Therefore, it is recommended that nursing managers pay attention to the level of emotional exhaustion among psychiatric nurses, implement effective support measures, and alleviate the impact of moral dilemma on turnover intention.Research has shown that while most psychiatric nurses have strong clinical skills and professional competence, they lack effective emotional management strategies and psychological support, which are key factors influencing their emotional exhaustion levels [30].It is recommended that nursing managers offer targeted emotional management training, establish psychological support systems, and improve the work environment to reduce nurses’ workload, thus helping them better handle work-related challenges, reduce emotional exhaustion, and decrease turnover intention [31].However, emotional exhaustion acts as an incomplete mediator in the relationship between moral dilemmas and turnover intention among psychiatric nurses.Therefore, additional research is needed to explore other factors influencing the relationship between moral dilemmas and turnover intention among psychiatric nurses, providing a theoretical basis for reducing turnover intention in this group.

### Study limitations

This study was limited to psychiatric nurses from four secondary and higher-level hospitals in Guangdong Province,and the results cannot be generalized as they rely on selfreporting. The findings of this study can only be generalized to its participants. Also, since it is a cross-sectional study, a cause–effect relationship cannot be established.

## Conclusions

The turnover intention among psychiatric nurses is at a moderately low level, with emotional exhaustion and moral dilemma being significant factors influencing turnover intention. Furthermore, emotional exhaustion plays a partial mediating role between moral dilemma and turnover intention.This suggests that management should focus on evaluating and preventing moral dilemmas among psychiatric nurses, while taking steps to reduce emotional exhaustion through supportive measures. This would mitigate the negative impact of moral dilemmas on turnover intention and help maintain the stability of the psychiatric nursing workforce.Future research could include multi-center, large-scale studies to further validate and enhance the findings of this study.Additionally, future research should consider other mediating or moderating variables and further explore the impact of moral dilemmas on turnover intention.

## Data Availability

All relevant data are within the manuscript and its Supporting Information files.

## Supporting information

**S1 Fig. Structural equation modeling of the relationships among Moral dilemma, emotional exhaustion, and turnover intention in psychiatric nurses**.Moral distress (latent construct derived from moral dilemma dimensions) is indicated by Insufficient autonomy, Failure to uphold patients’ best interests, and Value conflict (residuals e1– e3). Residuals for Emotional exhaustion and Turnover intention are e4 and e5.

Numbers on arrows are standardized coefficients: Moral distress → Emotional exhaustion *β* = 0.41; Emotional exhaustion → Turnover intention *β* = 0.42; Moral distress → Turnover intention β = 0.23; loadings to Moral distress = 0.96, 0.91, 0.81. All paths *p* < .01.Indirect effect of moral distress on turnover intention via emotional exhaustion = 0.172 (42.86% of total effect).

**S1 Dataset.Item-level responses and computed scores for Moral Distress and Emotional Exhaustion among healthcare workers, with demographics**

The dataset includes socio-demographic characteristics and psychological scale items. Socio-demographic variables were coded as follows: Gender (1=Male, 2=Female); Age (1=20–25, 2=26–30, 3=31–40, 4=>40); Marital status (1=Unmarried, 2=Married, 3=Divorced/Widowed); Professional title (1=Registered Nurse, 2=Nurse Practitioner, 3=Charge Nurse, 4=Assistant Head Nurse/Nurse Manager); Years of work experience (1=1–5, 2=6–10, 3=10–20, 4=>21); Education level (1=Associate or below, 2=Bachelor’s, 3=Master’s or above); Position (1=Clinical nurse, 2=Team leader, 3=Manager); Hospital grade (1=Secondary, 2=Tertiary); Employment method (1=Compilation, 2=Contract employment, 3=Labour dispatch); and Average monthly income (1=<5000 RMB, 2=5000–8000, 3=8000–12000, 4=>12000). Items A1–A20 represent the frequency of occurrence of moral dilemmas, rated 0–4 (0=Never, 4=Always); items B1–B20 represent the degree of distress for each dilemma, rated 0– 4 (0=Not at all distressing, 4=Extremely distressing). Items QX1–QX5 assess emotional exhaustion on a 0–6 scale, with higher scores indicating greater exhaustion. Item CZ measures turnover intention on a 1–5 scale, with higher scores reflecting stronger intention to leave.

## Acknowledgements

The authors thank all the participants in this study.

## Author Contributions

**Conceptualization:** Jiehui Yang,Rong Zhang,Yunling Zhong,Yana Wang,Shengyu Liu.

**Data curation:** Jiehui Yang.

**Formal analysis:** Yunling Zhong.

**Funding acquisition:** Rong Zhang.

**Investigation:** Jiehui Yang.

**Methodology:** Jiehui Yang,Rong Zhang,Yunling Zhong,Yana Wang,Shengyu Liu.

**Project administration:** Jiehui Yang.

**Resources:** Rong Zhang.

**Software:** Jiehui Yang.

**Supervision:** Rong Zhang.

**Validation:** Jiehui Yang,Rong Zhang,Yunling Zhong,Yana Wang,Shengyu Liu.

**Writing – original draft:** Jiehui Yang,Yana Wang,Shengyu Liu.

**Writing – review & editing:** Jiehui Yang,Rong Zhang,Yunling Zhong,Yana Wang,Shengyu Liu.

## References

1. Elangovan AR, Kar A, Steinke C. Meaningful moves: A meaning-based view of nurses’ turnover. Health Serv Manage Res. 2022;35(1):48–56. doi: 10.1177/09514848211010427. PMCID: PMC8795227.

2. Xu G, Zeng X, Wu X. Global prevalence of turnover intention among intensive care nurses: A meta-analysis. Nurs Crit Care. 2023;28(2):159–166. doi: 10.1111/nicc.12679. PMCID: 34261191.

3. Dai Z, Ma W. Modelling of Influencing Factors of Chinese Nurses’ Turnover Intention:A Meta-Analysis. Journal of PLA Nursing. 2021;38(8):5-7,15. Chinese.

4. Han X, Jiang F, Shen L, Liu Y, Liu T, Liu H, et al. Workplace Violence, Workforce Stability, and Well-being in China’s Psychiatric Hospitals. Am J Prev Med. 2022 Apr;62(4):e265–e273. doi: 10.1016/j.amepre.2021.09.013. PMCID: 34865934.

5. Luo Y, Bao J, Chen M,Mao C,Jia F,Yu Q, et al. The Correlation Analysis of Turnover Intention,Moral Distress and Stressor in Nurses. Chinese journal of industrial hygiene and occupational diseases. 2017; 36(8), 590-593.Chinese.

6. Wang XX, Wang LP, Wang QQ, Fang YY, Lv WJ, Huang HL, et al. Related factors influencing Chinese psychiatric nurses’ turnover: A cross-sectional study. J Psychiatr Ment Health Nurs. 2022 ;29(5):698–708. doi: 10.1111/jpm.12852. PMCID: 35716343.

7. Zhao BS, Wang J, Wang ML, Shi JJ D. J, Li FX. Meta-synthesis of qualitative studies on the authentic experience of moral dilemmas of ICU nurses. Chinese Journal of Nursing.2024;59(10), 1263–1269.Chinese.

8. Lamoureux S, Mitchell AE, Forster EM. Moral distress among acute mental health nurses: A systematic review. Nurs Ethics. 2024 ;31(7):1178–1195. doi: 10.1177/09697330241238337. PMCID: 38490947.

9. Wu M, Mao J, Dong QJ. Research on the Influence of Emotional Intelligence on the Turnover Intention of Special Education Teachers: The Chain Mediating Role Based on Emotional Labor and Emotional Exhaustion. Chinese Journal of Special Education.2023;(08), 76–84.Chinese.

10. Gehri B, Bachnick S, Schwendimann R, Simon M. Work-schedule management in psychiatric hospitals and its associations with nurses’ emotional exhaustion and intention to leave: A cross-sectional multicenter study. Int J Nurs Stud. 2023;146:104583. doi: 10.1016/j.ijnurstu.2023.104583. PMCID: 37619391.

11. Cao X,Qu JJ. Analysis of Origins and Main Contents of Conservation of Resource Theory and Implications. Human resources development of China.2014;(15), 75– 80.doi:10.16471/j.cnki.11-2822/c.2014.15.012.Chinese.

12. Zhang X, Huang H, Zhao S, Li D, Du H. Emotional exhaustion and turnover intentions among young ICU nurses: a model based on the job demands-resources theory. BMC Nurs. 2025;24(1):136. doi: 10.1186/s12912-025-02765-y. PMCID: 39915810.

13. Fumis RRL, Junqueira Amarante GA, de Fátima Nascimento A, Vieira Junior JM. Moral distress and its contribution to the development of burnout syndrome among critical care providers. Ann Intensive Care. 2017;7(1):71. doi: 10.1186/s13613-017-0293-2. PMCID: PMC5479870.

14. Maunder RG, Heeney ND, Greenberg RA, Jeffs LP, Wiesenfeld LA, Johnstone J, et al. The relationship between moral distress, burnout, and considering leaving a hospital job during the COVID-19 pandemic: a longitudinal survey. BMC Nurs. 2023 ;22(1):243. doi: 10.1186/s12912-023-01407-5. PMCID: PMC10369708.

15. Wu ML. Structural equation modeling: AMOS operation and application (pp. 52–53). Chongqing: Chongqing University Press; 2010.

16. Kendall MG, Stuart A, Ord JK. Kendall’s advanced theory of statistics. 6th ed. London: Edward Arnold; 1994.

17. Hamaideh S, Abu Khait A, Al-Modallal H, Masa’deh R, Hamdan-Mansour A, AlBashtawy M. Professional Quality of Life, Job Satisfaction, and Intention to Leave among Psychiatric Nurses: A Cross-Sectional Study. Nurs Rep. 2024 ;14(2):719–732. doi: 10.3390/nursrep14020055. PMCID: PMC11036228.

18. Liu XP. The compilation and preliminary application of Moral Distress Scale for psychiatric nurses. Central South University. 2023. Available from: https://kns.cnki.net/kcms2/article/abstract?v=3-fZNjprSazOeNayaI3GclSmxL2DGMh4q6VAbx0KwhIaPIYcHusmXMgMm8v_3XM8e_ggN4W3TBnjYhjcs7fdHrX wQ8vIJr72xqTgLvo83RT8wDa2UNVMFgrP-qnq4Vh4zgdZh-Z3kaWbCYDrtcIUrkplSCSkT6rwSTtEQUcgd28VvTGiYCfEqU_b7CO6WeMd&uniplatform=NZKPT&language=CHS

19. Li CP, Shi K. The influence of distributive justice and procedural justice on job burnout. Acta Psychologica Sinica.2003;35(5), 677– 684.doi:10.3724/SP.J.1041.2003.00677.Chinese.

20. Spector PE, Dwyer DJ, Jex SM. Behavior in organizations as a function of employee withdrawal: an empirical analysis of the turnover-intention relationship. J Appl Psychol. 1988;73(1):44–50.

21. Khaghanizadeh M, Koohi A, Ebadi A, Vahedian-Azimi A. The effect and comparison of training in ethical decision-making through lectures and group discussions on moral reasoning, moral distress and moral sensitivity in nurses: a clinical randomized controlled trial. BMC Med Ethics. 2023;24(1):58. doi: 10.1186/s12910-023-00938-5. PMCID: PMC10403849.

22. Xue B, Wang S, Chen D, Hu Z, Feng Y, Luo H. Moral distress, psychological capital, and burnout in registered nurses. Nurs Ethics. 2024 ;31(2-3):388–400. doi: 10.1177/09697330231202233. PMCID: 37737144.

23. He XS, Yang F, Liu LX,Cao S,Wang XY. The mediating effect of emotional exhaustion on the relationship between clinical nurses’ presenteeism and turnover intention. Journal of nursing administration. 2024;24(08), 657–661.Chinese.

24. Du XY, Mi JR, Yilixati AL, Liu YL. A Study on the Current Status and Influencing factors of Nurses’ Emotional Exhaustion at Tertiary Public Hospitals in China. Chinese Hospital Management.2024; 44(08), 23–28.Chinese.

25. Skaalvik EM, Skaalvik S. Job demands and job resources as predictors of teacher motivation and well-being. Soc Psychol Educ. 2018;21(5):1251–1275. doi:10.1007/s11218-018-9464-8.

26. He XC, Zhou WQ, Zhang BL, Xia ZC,Wei YH,He XJ,et al. Current situation and influencing factors of turnover intention of psychiatric nurses. Chinese evidence-based nursing.2023;9(12), 2253–2258.Chinese.

27. Liu XC, Ma YZ, Zhang Y. Influence of Satir model therapy on stress-coping styles, psychological resilience and self-efficacy of re-employed nurses. Chinese nursing management.2023; 23(07), 1048–1053.Chinese.

28. Meng JT, Zhou L, Xia XL,Xu XB,Cao WL, Liu Y. Research progress of moral injury in clinical nursing. Journal of nursing science.2024;39(05), 116–120.Chinese.

29. Wu XQ, Yang Z, Xie ZQ, D. YY,Yang LN,Chen SH. Summary of the best evidence for nurses moral dilemma intervention. Journal of nursing(China).2023;30(01), 47–52.Chinese.

30. Li LZ, Yang P, Singer SJ, Pfeffer J, Mathur MB, Shanafelt T. Nurse Burnout and Patient Safety, Satisfaction, and Quality of Care: A Systematic Review and Meta-Analysis. JAMA Netw Open. 2024 ;7(11):e2443059. doi: 10.1001/jamanetworkopen.2024.43059. PMCID: PMC11539016.

31. Lee M, Cha C. Interventions to reduce burnout among clinical nurses: systematic review and meta-analysis. Sci Rep. 2023;13(1):10971. doi: 10.1038/s41598-023-38169-8. PMCID: PMC10325963.

